# Incidence of COVID-19 reinfection: an analysis of outpatient-based data in the United States of America

**DOI:** 10.1101/2021.12.07.21267206

**Authors:** Mahalul Azam, Feddy Setio Pribadi, Arief Rahadian, Muhammad Zakki Saefurrohim, Yudhy Dharmawan, Arulita Ika Fibriana, Ima Arum Lestarini, Syed Mohamed Aljunid

## Abstract

**Objectives:** COVID-19 reinfection cases are evidence of antibody waning in recovered individuals. Previous studies had reported cases of COVID-19 reinfection both in hospital-based and community-based data. However, limited studies reported COVID-19 reinfection in large community-based data. The present study aimed to provide the incidence of COVID-19 reinfection based on secondary data in the U.S.

**Study design:** Cross-sectional study

**Methods:** A cross-sectional study was conducted using secondary data provided by COVID-19 Research Database, i.e., Healthjump. Reinfection were defined as diagnosed COVID-19 (U07.1= confirmed virus identified) twice with ≥90 days interval between diagnosis. Age, gender, and region data were also explored. A Chi-square test continued by a binary logistic regression was conducted to determine the association between parameters. Data collecting and processing were done in the Amazon workspace.

**Results:** The study revealed 3,778 reinfection cases of 116,932 COVID-19 infected cases (3.23%). Reinfection cases were more common in females (3.35%) than males (3.23%). Elderly subjects were the highest incidence (5.13%), followed by adult (4.14%), young adults (2.35%), and children (1.09%). Proportion in the region of living northeast was the highest (3.68%), compared to the south (3.49%), west (2.59%), and midwest (2.48%).

**Conclusion:** The incidence of COVID-19 reinfection was 3.23%, suggesting our concern with COVID-19 management and future research to understand COVID-19 reinfection better. The incident is more likely to occur in female and elderly patients.

## 1. Introduction

Coronavirus diseases-19 (COVID-19) has infected more than 216 million people and almost 4.5 million deaths since first found in Wuhan.(1) Individuals who have recovered from COVID-19 are protected from future SARS-CoV-2 infection by producing specific antibodies.(2) Some reports suggested that primary infection can provide better protection against reinfection in the health care population and the general population.(2,3) Previous studies reported naive infection to provide better protection from reinfection compared to naive vaccination,(3) some other studies reported on the contrary.(4–6).

On the other hand, some studies recently reported cases of COVID-19 reinfection both in hospital-based and community-based data.(7–12) Reinfection cases are the evidence of antibody waning or loss of recovered individuals.(13) It is crucial to understand more about reinfection incidence and its characteristics to mitigate the COVID-19 pandemic better. Limited studies provide evidence of COVID-19 reinfection in extensive community-based data. The present study aimed to provide the incidence of COVID-19 reinfection based on secondary data in the U.S.

## 2. Methods

A cross-sectional study was conducted to explore the incidence of COVID-19 reinfection in outpatient recorded secondary data. The primary endpoint to be observed is the occurrence of COVID-19 reinfection. We also observed determinant factors of COVID-19 reinfection, such as a patient’s characteristics and demography.

### 2.1 Variable determination

COVID-19 reinfection were determined as the second positive result of reverse transcription-polymerase chain reaction (RT-PCR) after the first positive result that was diagnosed more or equal to 90 days.(14,15) A positive result of RT-PCR data was provided in the diagnosis data as confirmed COVID-19 (code U07.1). To determine COVID-19 reinfection at different times; the encounter data of outpatients are needed (as stated in the data set). We determined the interval between the first and second diagnosis COVID-19 for more or equal to 90 based on the Centers for Disease Control and Prevention (C.D.C.) protocol for suspected reinfection cases. The patient’s characteristics consist of age, gender, and region. Age was categorized as child for subjects with 0-14 years old, young for subjects aged 15-47 years old, adult for 48-63 years old, and elderly for 64 years old or above. The region was categorized into four main regions in the U.S., i.e., northeast, midwest, south, and west as published by U.S. Census Bureau at https://www2.census.gov/geo/pdfs/maps-data/maps/reference/us_regdiv.pdf

### 2.2 Data collection

We used secondary data provided by COVID-19 Research Database (https://covid19researchdatabase.org/), i.e., Healthjump.

The Healthjump application exports data for the electronic health records (E.H.R.) & practice management (PM) system, providing all data that was changed from the last time the application ran. It is up to the receiving application to append this data to the data previously received. The data involved all regions and states in the U.S.

### 2.3 Identifying variables and parameters

The study population of this study were all subjects in outpatients diagnosed as COVID-19; we then determined the COVID-19 reinfection incidence in the study population as stated in the variable determination, i.e., diagnosed COVID-19 twice with a 90 days period between diagnosis. The period of data capturing be noticed in the data analysis.

Determination of data are as follows:

– Subjects with COVID-19 determined from Healthjump data source with the ICD-10 diagnosis code U07.1 (confirmed virus identified).
– COVID-19 reinfection cases were extracted from the diagnosis data in the Healthjump data source. To determine this status, subjects should have two times identified in the diagnosis code with U07.1, i.e., diagnosed as confirmed COVID-19 in different encounter id and the range between encounter dates should be ≥ 90 days. The period was determined from the diagnosis date provided in the diagnosis tables in the Healthjump data source.
– We also accessed the Healthjump data source for age (calculated from date of birth (DoB) and encounter_date), gender, and regions that converted from states provided in the addresses.
– After all needed data was extracted, we then cleaned the data to ensure no missing data in the data set. Data cleaning were conducted by eliminating incomplete, missing values, or not meeting the requirements. Missing data analysis were conducted in this section. Data then were processed to present the findings and conclude the pivotal results.

### 2.4 Statistical Analysis

The clean and firmed data had been analyzed and presented based on the type of data—nominal data presented in proportion. Analysis to determine the association between parameters and primary outcomes was conducted. Binary logistic regression has been conducted to conclude the regression model related to the occurrence of COVID-19 reinfection. Before building a binary logistic regression model, the confounding analysis, the interaction effect of the determinants, the nonlinear data effect, and the goodness of fit were considered. We developed a regression model of the particular parameters to be visualized. Statistical analysis as well as general data analysis was performed using STATA that was available in the workspace.

### 2.5 Working in the Workspace Environment

We conducted our study on Snowflake using Python to process data from the provided Covid 19 Research Database. We choose Python as our tool because there are available methods that have been provided, especially in machine learning. The display in Python also helped us do data analysis because Python can display the program output in one frame. Our team is also very familiar with the Python programming language, so it was easy to understand the program codes included in the workspace guide.

## 3. Ethical Approval

The COVID-19 Research Database was established with institutional review board/ patients advocacy and ethics approval (https://covid19researchdatabase.org/) and exemption from patient consent due to the use of Health Insurance Portability and Accountability Act (HIPAA) de-identified data, HIPAA limited data, or non-HIPAA covered data, along with strong governance measures in place to control access to all data. This exemption covers all research performed within the COVID-19 Research Database. COVID-19 Research Database Statement of Ethics and Intent provided at https://covid19researchdatabase.org/statement-of-ethics/

## 4. Result

Our study acquired 195,911 COVID-19 confirmed cases with the diagnosis code U07.1. Of them, 78,979 had incomplete data, and finally, 116,932 were included in the analysis. (Figure 1) The present study revealed that the incidence of COVID-19 reinfection cases was 3.23 % (3,778) from 116,932 infected people in our data (Table 1). Table 1 also showed that COVID-19 infection in female was significantly more common than male (56.88 % vs 43.12%) and so did the COVID-19 reinfection (2,225 (3.35%) vs 1,553 (3.23%)) (Table 2). In the age category, the young adult was the most common, i.e., 55.12% (64,449 cases) who got COVID-19 infection, but COVID-19 reinfection occurred more common in the elderly (1,355 or 5.13%). South America had the highest number of COVID-19 infections, i.e., 67,670 (57.87%); however, the highest percentage of COVID-19 reinfections was in the northeast region as 3.68% or 496 cases and the south region placed in the second highest as 3.49% or 2,363 cases.

**Table 1.** Subjects’ characteristics.

| No | Characteristics | Total subjects: 116,932 |  |
| --- | --- | --- | --- |
|  |  | N | % |
| 1 | Reinfection |  |  |
|  | Yes | 3,778 | 3.23 |
|  | No | 113,154 | 96.77 |
| 2 | Sex |  |  |
|  | Female | 66,511 | 56.88 |
|  | Male | 50,421 | 43.12 |
| 3 | Age |  |  |
|  | Child | 5,601 | 4.79 |
|  | Young adult | 64,449 | 55.12 |
|  | Adult | 20,447 | 17.49 |
|  | Elderly | 26,435 | 22.61 |
| 4 | Region |  |  |
|  | Northeast | 13,461 | 11.51 |
|  | Midwest | 7,267 | 6.21 |
|  | South | 67,670 | 57.87 |
|  | West | 28,534 | 24.40 |

**Table 2.** Subjects’ characteristics based on Reinfection status.

| No | Characteristics | COVID-19 Reinfection |  |  |  | Total | P* |
| --- | --- | --- | --- | --- | --- | --- | --- |
|  |  | Yes |  | No |  |  |  |
|  |  | N<br>(3,778) | % | N<br>(113,154) | % |  |  |
| 1 | Sex |  |  |  |  |  | 0.0001 |
|  | Female | 2,225 | 3.35 | 64,286 | 96.65 | 66,511 |  |
|  | Male | 1,553 | 3.23 | 48,868 | 96.92 | 50,421 |  |
| 2 | Age |  |  |  |  |  | 0.0001 |
|  | Child | 61 | 1.09 | 5,540 | 98.91 | 5,601 |  |
|  | Young adult | 1,515 | 2.35 | 62,934 | 97.65 | 64,449 |  |
|  | Adult | 847 | 4.14 | 19,600 | 95.86 | 20,447 |  |
|  | Elderly | 1,355 | 5.13 | 25,080 | 94.87 | 26,435 |  |
| 3 | Region |  |  |  |  |  | 0.0001 |
|  | Northeast | 496 | 3.68 | 12,965 | 96.32 | 13,461 |  |
|  | Midwest | 180 | 2.48 | 7,087 | 97.52 | 7,267 |  |
|  | South | 2,363 | 3.49 | 65,307 | 96.51 | 67,670 |  |
|  | West | 739 | 2.59 | 27,795 | 97.41 | 28,534 |  |

**Figure 1.**
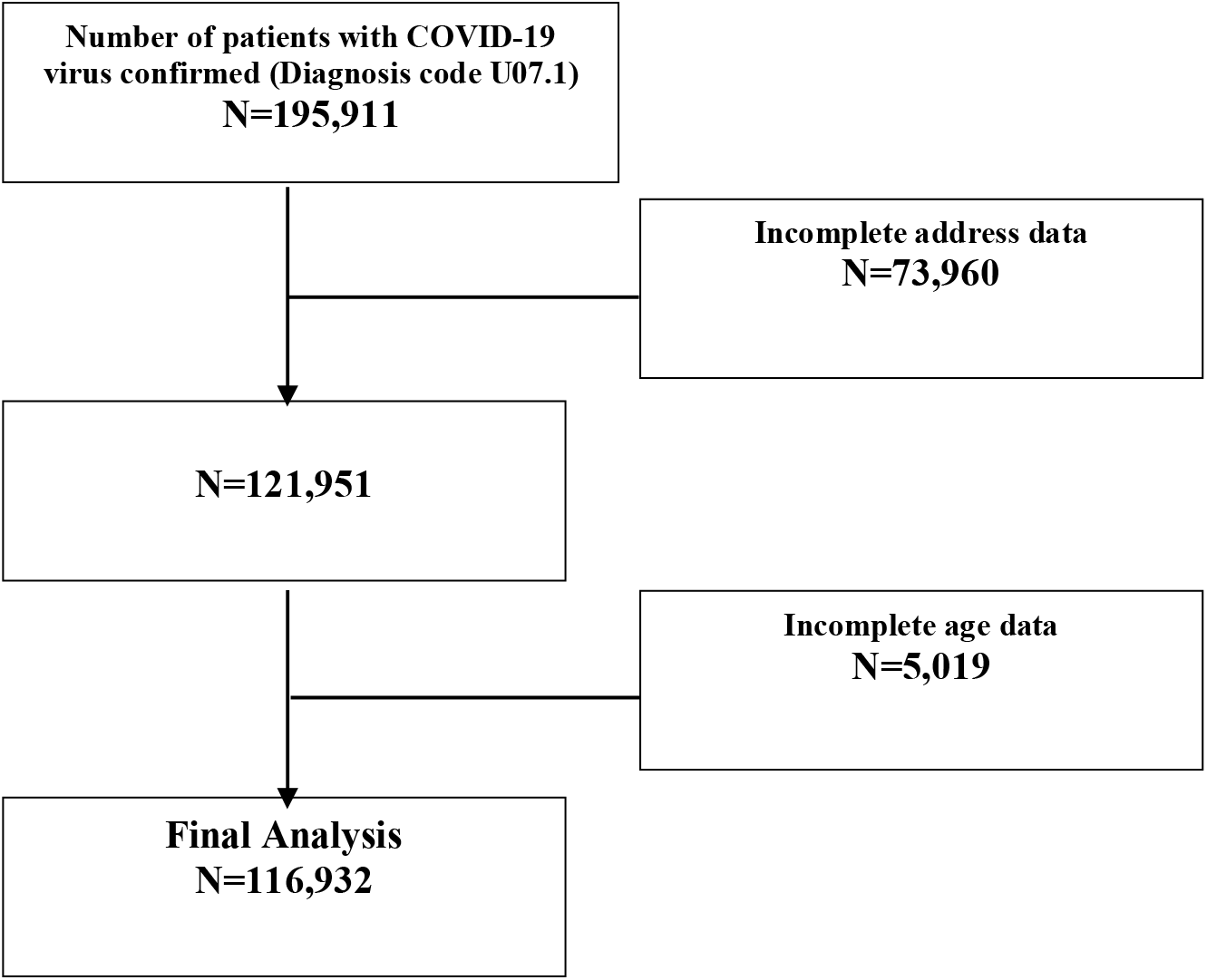
Subjects selection chart.

A binary logistic regression was then conducted and found that females, the elderly, and those living in the south region had a significantly higher risk of getting COVID-19 reinfection cases (Table 3). The coefficient estimate was -0.099 for males, which means the event of COVID-19 reinfection is a factor exp (−0.099) = 0.91 or 9 % lower in males than females when the group of age and region is equal. This binary logistic regression also showed us that the elderly group living in the south region had a higher risk than others.

**Table 3.** Binary logistic regression of subjects’ characteristics for COVID-19 reinfection Standard.

|  | **Coeffisient** | **Standard Error** | **z** | **P>[Z]** | **95% C.I.** |  |
| --- | --- | --- | --- | --- | --- | --- |
| Male | -0,099 | 0,033 | -2,96 | 0,003 | -0,165 | -0,033 |
| Young | 0,766 | 1,131 | 5,83 | <0,001 | 0,509 | 1,024 |
| Adult | 1,339 | 0,133 | 10,03 | <0,001 | 1,078 | 1,601 |
| Elderly | 1,557 | 0,131 | 11,81 | <0,001 | 1,298 | 1,815 |
| Midwest | -0,396 | 0,088 | -4,49 | <0,001 | -0,570 | -0,223 |
| South | -0,066 | 0,050 | -1,33 | 0,185 | -0,165 | 0,031 |
| West | -0,263 | 0,059 | -4,44 | <0,001 | -0,379 | -0,147 |
| Constant | -4,324 | 0,137 | -31,41 | <0,001 | -4,593 | -4,054 |

## 5. Discussion

The present study reported the incidence of COVID-19 reinfection, i.e., 3.23% of total 116,932 infected cases, based on the U.S. community-based data acquired from outpatients’ secondary data recorded on the Healthjump database. This finding adds the evidence of COVID-19 reinfection based on large community data, which is limited studies have been reported. Compared with previous reports, the incidence of COVID-19 reinfection in this study was higher. The previous community-based study was also conducted in Austria and revealed that reinfection incidence was 0.27% of 14,840 cases.(10) Previous study used total national records of COVID-19 infection data from the Austrian epidemiological reporting system. Reinfection was considered for patients who had a positive polymerase chain reaction (PCR) test during the first and second infection wave. The first wave was from February to April 2020, while the second wave was from September to November 2020, which means that the reinfection interval ranged from 5 months to 9 months. Our study determined that the interval period was equal or more than 90 days (3 months). Another study by Slezak et al. (16), reported a COVID-19 reinfection incidence rate of 0.8% from 75,149 total cases. Slezak’s study defined the term of reinfection equal to the current study, i.e., a positive PCR test for SARS-CoV-2 ≥ 90 days after the first positive test. However, the Slezak study conducted a sensitivity analysis using a suspected reinfection definition of ≥ 120 days to anticipate a potential late viral shedding that might influence the result.(16) Previous study in Qatar revealed the incidence of COVID-19 reinfection was 0.18% from 133,266 cases.(17) Qatar study categorized twice detected PCR test as (1) suspected reinfection for patients with the interval period of ≥ 45 days; (2) strong evidence reinfection for patients meets category one and had Ct value of <30; good evidence reinfection for patients with Ct values ≥ 30 and was associated with the supporting contextual evidence: the appearance of symptoms and closed-contact history.(17) A systematic review and meta-analysis also reported that the pooled estimated incidence of COVID-19 reinfection was 0.3%.(18) The determinations of reinfection each study in this systematic review were varied, i.e., based on interval period only or viral genome sequencing confirmation.(18)

Determination of the interval period between the first and second infection related to the proportion of COVID-19 reinfection incidence. Previous studies which were not considered a recurrence positivity period equal to or more than 90 days reported a higher incidence rate. Our previous systematic review study revealed that 14.8% of recovered COVID-19 patients acquired re-positivity after negative test results.(9) Our previous review did not consider the interval period between both infections. Another previous review conducted by Dao et al. (19) revealed that re-positivity proportions were varied from 2.4 % to 69.2 %. The potential cause of re-positivity in this study might also be varied, such as false positive which is used in the first or second diagnosis, false negatives, which were used to determine recovery, reactivation or were reinfected with another SARS-CoV-2 strain.(19)

In the general term, reinfection was determined that a person was infected with an agent, recovered, and became infected again.(18) Present study determined COVID-19 reinfection as persons with at least one detection of SARS-Cov□2 R.N.A. test, equal or more than 90 days after the first detection of SARS-Cov□2 R.N.A., whether or not symptoms were present.(14,15,18) Yahav et al. describe reinfection can be any positive R.T.-PCR more than 90 days since the first episode. However, Yahav et al. also explained the need to know Ct value < 35 to conclude COVID-19 infection, yet explaining any suspect of reinfection needs to be considered isolation.(14) Persons with COVID□19□like symptoms and SARS□Cov□2 R.N.A. detection between 45 and 89 days since first SARS□CoV□2 infection is still considered COVID-19 reinfection. This condition is supported by the evidence of close□contacts and without evidence of another cause of infections.(18) In the present study, data regarding close contacts were not provided in the database. For epidemiological confirmation, viral genotype assays of the first and second specimens are needed to conclude two different phylogenetic strains by high-throughput sequencing, corresponding to local epidemiology (proof of two episodes with two distinct virus variants with any sequence variation);(14,18,20,21) For clinical practice: reinfection may be defined as clinical recurrence of symptoms compatible with COVID-19, accompanied by positive PCR test (Ct < 35), more than 90 days after the onset of the primary infection, supported by close-contact exposure or outbreak settings, and no evidence of another cause of infection. In the presence of epidemiological risk factors (i.e., significant exposure), reinfection should be considered during the first 90 days if clinical symptoms of the first episode resolved and two PCR tests were negative before the new episode. Viral culture, if collected, is expected to be positive. (14,18,20,21) Likewise, epidemiological and clinical practice data were also not provided in the Healthjump database.

The present study adds COVID-19 reinfection evidence defined as cases that are twice diagnosis COVID-19 (U07.1), with the interval between diagnosis ≥ 90 days. Although prior infection protects against reinfection(22), reinfection cases are still revealed in this study. Indeed, other studies also concluded that reinfection occurred with mild symptoms and faster recovery. (23,24) Previous studies also reported that the estimated antibody levels associated with protection against reinfection likely last 1.5-2 years on average, with levels related to protection from severe infection present for several years. (25) Another study reported that rapid waning of COVID-19 antibody responses attenuated after 90 days;(26) however, a recent longer follow-up study showed that antibody titers remained stable over four months(27), and this presence of antibody actually was not equal to the protective immunity.(26)

The current study also concluded that several factors may affect the occurrence of reinfection and revealed that sex was a significant factor affecting reinfection, where females are more common than males. The association between gender and risk of reinfection is still a contradiction. A recent systematic review by Ghorbani et al. stated that males have a higher reinfection rate than females.(18) However, another study(7,16) concluded in line with the current study which females were more common to be reinfected than males. Another study(8) concluded no difference between sex and the occurrence of COVID-19 reinfection. This study also deduced that reinfection was more common in the elderly (>64 years old) compared to the other age categories. A recent systematic review stated that reinfection occurred in patients aged 15 to 99 years old.(18) Another previous study stated that the average age of the reinfection cases was 50 years old.(11) Study in Southern California concluded that COVID-19 reinfections were common in the elderly, and this study also concluded that reinfections were related to the immunocompromised patients.(16)

The present study also revealed that the highest COVID-19 reinfection cases proportion was in the northeast region (3.68%), followed by the south region (3.49%), west region (2.59%), and midwest region (2.48%). This proportion was not in line with the proportion of infection cases, which the highest was the south region (67.67%), west region (28.53%), northeast (13.46%), and midwest region (7.27%). The reinfection proportion was also not in line with the proportion of the population within the regions, i.e., northeast region (49.29%), midwest region (31.81%), south region (17.13%), and west region (1.77%).(28)

The next important step for future mitigation of COVID-19 is whether reinfection will represent a serious problem or not. The case of reinfection in COVID-19 is still elusive. Many cases of reinfection have been reported from all over the world. The limitation of our study is that our data did not include genome sequencing data of the virus so that we could not differentiate between reinfection or reactivation of the virus. Nevertheless, the three months difference between each PCR test provided in the diagnosis data has been accepted as a period that is possible for reinfection because of the reduction of antibodies, yet the least possible for reactivation.

## 6. Conclusion

The present study concluded that the incidence of COVID-19 reinfection is higher than previous studies, i.e., 3.23%, suggesting our concern on COVID-19 management and future research to understand COVID-19 reinfection better. The incident is more likely to occur in female and elderly patients.

## Data Availability

Data acquired are subject to sharing restrictions, and the authors are legally prohibited from sharing them. However, these data are available upon request from the Covid-19 Research Database Consortium. Data processing was done under the Covid-19 Research Database workspace environment. It should be noted that to protect patient privacy; the Covid-19 Research Database does not permit researchers to export record-level data. Requests to access this data should be sent to or via the Submission Hub at https://covid19researchdatabase.org.

## Acknowledgment

The data, technology, and services used in the generation of these research findings were supplied pro bono by the COVID-19 Research Database partners, who are acknowledged at https://covid19researchdatabase.org.

## Author Contributions

Conceptualization, M.A.; methodology, M.A. and A.R.; formal analysis, M.A. and Y.D.; investigation, M.A., M.Z.S, and F.S.P.; data curation, F.S.P, and Y.D.; writing—original draft preparation, M.A. and A.R..; writing—review and editing, M.A., A.R. and A.I.F; supervision, A.I.F, I.A.L, and S.M.A. All authors have read and agreed to the published version of the manuscript.

